# Maternal first trimester metabolic profile in TGA pregnancies - a case-control study

**DOI:** 10.1101/2022.01.11.22268842

**Authors:** Johanna Huida, Tiina Ojala, Johanna Hautala, Heljä-Marja Surcel, James R Priest, Emmi Helle

## Abstract

**Objective:** To evaluate the association between maternal first trimester metabolic profile and transposition of the great arteries (TGA) in the offspring.

**Design:** A matched case-control study using national register data and maternal first trimester blood samples.

**Settings:** Finland

**Patients:** A total of 100 mothers with TGA in their offspring and 200 controls matched by the year of birth and sex of the child, and the age and the body mass index (BMI) of the mother.

**Interventions:** None.

**Main outcome measures:** To identify the impact of 73 metabolic measure concentrations on the likelihood that a child would be born with TGA.

**Results:** A higher concentration of four subtypes in extremely large very-low-density lipoprotein (VLDL) particles and one in large VLDL particles were observed in mothers with TGA in their offspring. This finding did not reach statistical significance after multiple testing correction. In the analysis of the subgroups of maternal BMI over 25 and 30 only, the odds ratios (OR) of the metabolic variables were higher compared to the original population. In the group with a BMI over 25 the mean OR was 1.3 (N= 111, total) and in the group with a BMI over 30 the mean OR was 2.1 (N= 42, total).

**Conclusions:** We found no statistically significant difference between the metabolic profiles of mothers with TGA in their offspring, and their controls. However, the trend in our BMI-matched study potentially indicates that the likelihood of TGA development associates with maternal early-pregnancy metabolic profile, and the association is more pronounced in the subgroups of BMI over 25 and 30. Further studies are needed especially in the higher BMI groups.

**What is already known on this topic?:**

- The most important structures of the heart develop during early pregnancy, but studies on the association of maternal metabolomics during the first trimester with CHD development are scarce.
- Maternal obesity and diabetes are known risk factors for CHD in the offspring but the precise developmental mechanisms underlying this risk remain unknown.

**What this study adds to this topic?:**

- The maternal metabolomic profile in early pregnancy is potentially associated with the likelihood of TGA development in the offspring.
- Metabolic markers in obese women may have a stronger association with the development of TGA but this connection requires further studies.

## Introduction

Congenital heart defects (CHD) are the most common congenital anomalies, and transposition of the great arteries (TGA) is one of the most frequent cyanotic CHD in neonates. The birth prevalence of TGA is approximately 2–3 per 10,000 (1,2). Maternal diabetes (3,4), higher haemoglobin A1C (5), and elevated random plasma glucose values in the first trimester in mothers without diabetes (6) have been associated with an increased risk for CHD in the offspring. The risk is increased even in diabetic mothers with good glycaemic control (5,7). The prevalence of all types of diabetes has been increasing among women of fertile age (8). At the same time, the prevalence of obesity in women of childbearing age is increasing worldwide(9). Pre-pregnancy obesity is known to increase the risk for gestational diabetes mellitus (GDM) (10). Previous studies have further consistently reported maternal overweight as a risk factor for CHD in the offspring and particularly for TGA (3). This risk increases along with the severity of obesity (11–13).

Abnormal glucose metabolism does not, however, entirely explain the increased risk for CHD in the offspring of obese mothers (14). The causal relationship and increased risk are likely to be multifactorial including different cardiometabolic factors and epigenetic mechanisms. Only a few studies have previously assessed the metabolomic profile of maternal blood during the first trimester of pregnancy, which reflects the metabolic circumstances at the time of cardiac morphogenesis.

The aim of our study was to further elucidate the association between maternal metabolic risk factors and the risk for CHD in the offspring. We compared a panel of first-trimester metabolic markers among women who delivered a child with TGA and women who delivered a healthy child in a setting of a matched case-control study. Identifying new risk biomarkers could potentially enable prevention with public health recommendations for women planning pregnancy and help to direct resources into the screening of risk groups.

## Methods

All expectant mothers in Finland are entitled to a follow-up at an outpatient maternity clinic. In conjunction with their first visit to the clinic, the expectant mothers are screened for infectious diseases: syphilis, HIV and hepatitis B. These screening serum samples collected over the years 1983–2016 (Finnish Maternity Cohort) were deposited at Biobank Borealis if the mother consented to this (15).

From a national cohort of all live-born infants with simple d-TGA (TGA +/- small ventricular septal defect, n=127) born in Finland during 2004–2014 (16), we identified 100 live-born cases and each case was matched with two healthy controls (n=200) whose maternal screening samples were available at Biobank Borealis. The background data was collected from the Medical Birth Register maintained by the Finnish Institute for Health and Welfare. The cases and controls were matched according to the sex of the child, year of birth, and the age and the body mass index (BMI) of the mother. Maternal BMI was calculated from the register data based on maternal pre-pregnancy weight and height.

The maternal screening serum samples were requested from the Biobank Borealis and analysed using an automated high-throughput serum NMR metabolomics platform (17).

To describe the study population, the statistical methods were chosen based on the statistical features of the data (Table 1). Categorical variables were presented as percentages and frequencies. Continuous variables were described with medians and interquartile ranges (IQRs) or means and standard deviations depending on the normality of the variable. Birth weight, length and head circumference were measured in grams and centimetres, and then converted into SD units (i.e. deviation from Finnish general population mean accounting for plurality and parity (18)). Maternal smoking was defined as any smoking during pregnancy including those who ceased smoking during the first trimester of pregnancy.

**Table 1.**

| Characteristic | CHD (N= 99) | No CHD (N= 197) | P -value |
| --- | --- | --- | --- |
| Age, years (median, IQR) | 29 (27-34) (N=99) | 29 (27-33.5) (N=197) | NS * |
| BMI, kg /m2 (median, IQR) | 23.31 (21.01-27.01) (N=99) | 23.92 (21.55-27.08) (N=197) | NS * |
| Sex of the child |  |  |  |
| Boy | 74.7 % (N=74) | 74.6 % (N=147) | NS** |
| Girl | 25.3 % (N=25) | 25.4 % (N=50) |  |
| Parity |  |  |  |
| Nulliparous | 25.3 % (N=25) | 33.5 % (N = 66) | NS** |
| Multiparous | 74.7 % (N= 74) | 66.5 % (N = 131) |  |
| Smoking during pregnancy |  |  |  |
| Non-smoker | 82.5 % ( N= 80) | 85.9 % ( N= 164) | NS** |
| Smoker | 17.5 % (N= 17) | 14.1 % (N= 27 ) |  |
| Oral glucose tolerance test done |  |  |  |
| Yes | 39.4 % (N=39) | 43.1 % (N=85) | NS ** |
| Oral glucose tolerance test pathological if done |  |  |  |
| Yes | 28,2 % (N=11) | 24,7 % (N=21) | NS ** |
| Number of foetuses |  |  | NS** |
| 1 | 96 % (N=95) | 98.5 % (N=194) |  |
| 2 | 4% (N= 4) | 1.5 % (N=3) |  |
| Method of childbirth |  |  |  |
| Spontaneous vaginal birth | 74.7 % (N=74) | 75.1 % (N=148) | NS ** |
| Ventouse cup | 7.1 % (N=7) | 7.1 % (N=14) |  |
| Elective caesarean section | 4 % (N=4) | 6.6 % (N=13) |  |
| Urgent caesarean section | 13.1 % ( N=13) | 7.1 % (N=14) |  |
| Emergency caesarean section | 1 % (N= 1) | 2.5 % (N=5) |  |
| Vaginal breech birth | 0% (N=0) | 1.5 % (N=3) |  |
| Gestational Age, d (median, IQR) | 278 (270-284) (N=99) | 281 (275-286) (N=197) | 0.01 * |
| Birth weight, SD units (mean, SD) | -0.3113 (1.15991) (N=97) | -0,0604 (1.08452) (N= 197) | NS*** |
| Birth height, SD units (mean, SD) | -0.0971 (1.02660) (N=70) | -0,0589 (1.05428) (N=197) | NS*** |
| Head circumference, SD units (mean, SD) | -0.6032 (1.01690) (N=62) | 0,0735 (1.02922) (N=196) | 0.000 *** |

Patient background characteristics from national registers.
Values are mean values (standard deviation, SD) for normally distributed and median (interquartile range, IQR) for skewed variables.
\* Mann-Whitney U-test
\*\* Chi -Square Test of Independence
\*\*\* Independent samples T-Test
BMI, body mass index; NS, non significant

The metabolic measures are absolute concentration units (i.e., millimoles per litre) of different magnitudes. The normality of the distributions were analysed by visual inspection of histograms and using skewness as a test of normality. The measures were log-transformed when skewed (skewness > 1) and scaled to standardised values (Z-score) prior to analysis to allow comparison of associations across metabolites. We excluded one case and its controls and one control because of outliers exceeding the Z-score +/- 5 in more than one variable. 0-values were excluded.

Conditional logistic regression was performed to assess the impact of higher metabolic measure concentration on the likelihood that a mother would have a child with TGA. This analysis took into account the stratification and matching of the data. BMI was included as a covariate. Analysis was undertaken on 73 metabolic measures. Due to the correlated nature of the data, principal component analysis was used to evaluate the appropriate number of independent tests for the correction of multiple comparisons(19). In our analysis, principal component analysis revealed the presence of four components explaining 94.9 % of the variation and therefore p-value < 0.0125 (0.05/4) was used as a threshold for statistical significance.

All statistical analyses were conducted with SPSS version 27 (IBM Corp.).

The study protocol was reviewed and approved by the Ethics Committee of the Hospital District of Helsinki and Uusimaa (HUS).

## Results

The baseline characteristics of the study population are presented in Table 1. A total of 39 cases and 85 controls had undergone the 75 g oral glucose tolerance test (OGTT). Of these, the OGTT was pathological in 11 (28,2 %) of mothers with TGA children and in 21 (24,7 %) of the controls (p -value 1.0). The distribution of maternal BMI was similar in the two groups, and 38 % of expectant mothers were overweight or obese (Figure 1). The head circumference (HC) differed in the two groups as TGA infants had significantly lower HC in SD units adjusted for GA when compared to the controls (p-value < 0.01, CI -0.971- (- 0.382)) (Table 1).

**Figure 1.**
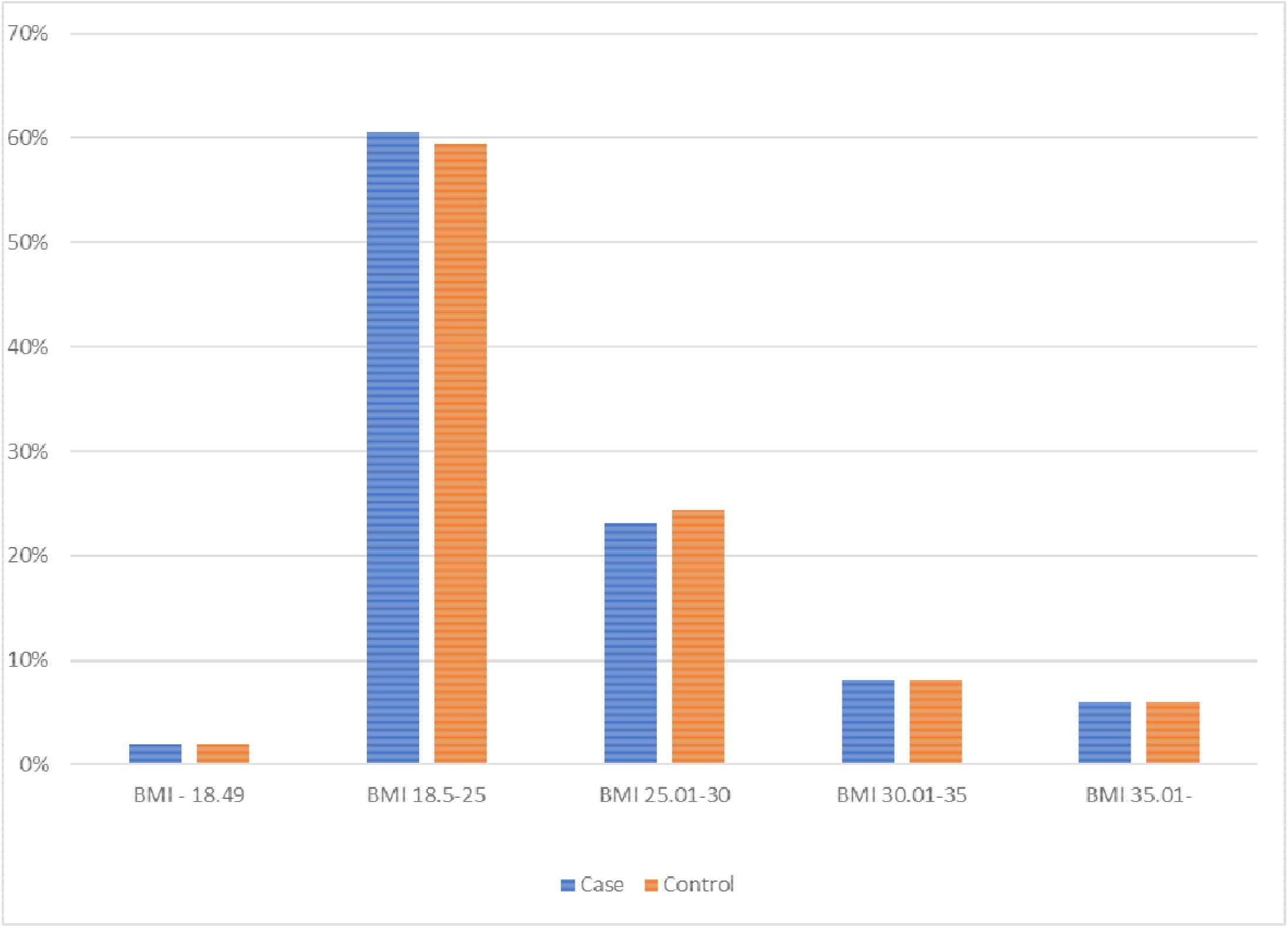
The BMI distribution between cases and controls in percentages (columns)

The metabolic marker values are presented in Figure 2. Five of the metabolic measures (concentration of chylomicrons and extremely large VLDL particles, total lipids in chylomicrons and extremely large VLDL, free cholesterol in chylomicrons and extremely large VLDL, triglycerides in chylomicrons and extremely large VLDL, free cholesterol in large VLDL) were found as predictors of TGA with a p-value < 0.05 in univariate analyses. However, after multiple testing correction they did not remain statistically significant (p-value < 0.0125). There was no statistically significant difference in metabolic markers explaining the smaller HC in TGA neonates and there was no correlation between HC and metabolic markers measured (data not shown).

**Figure 2.**
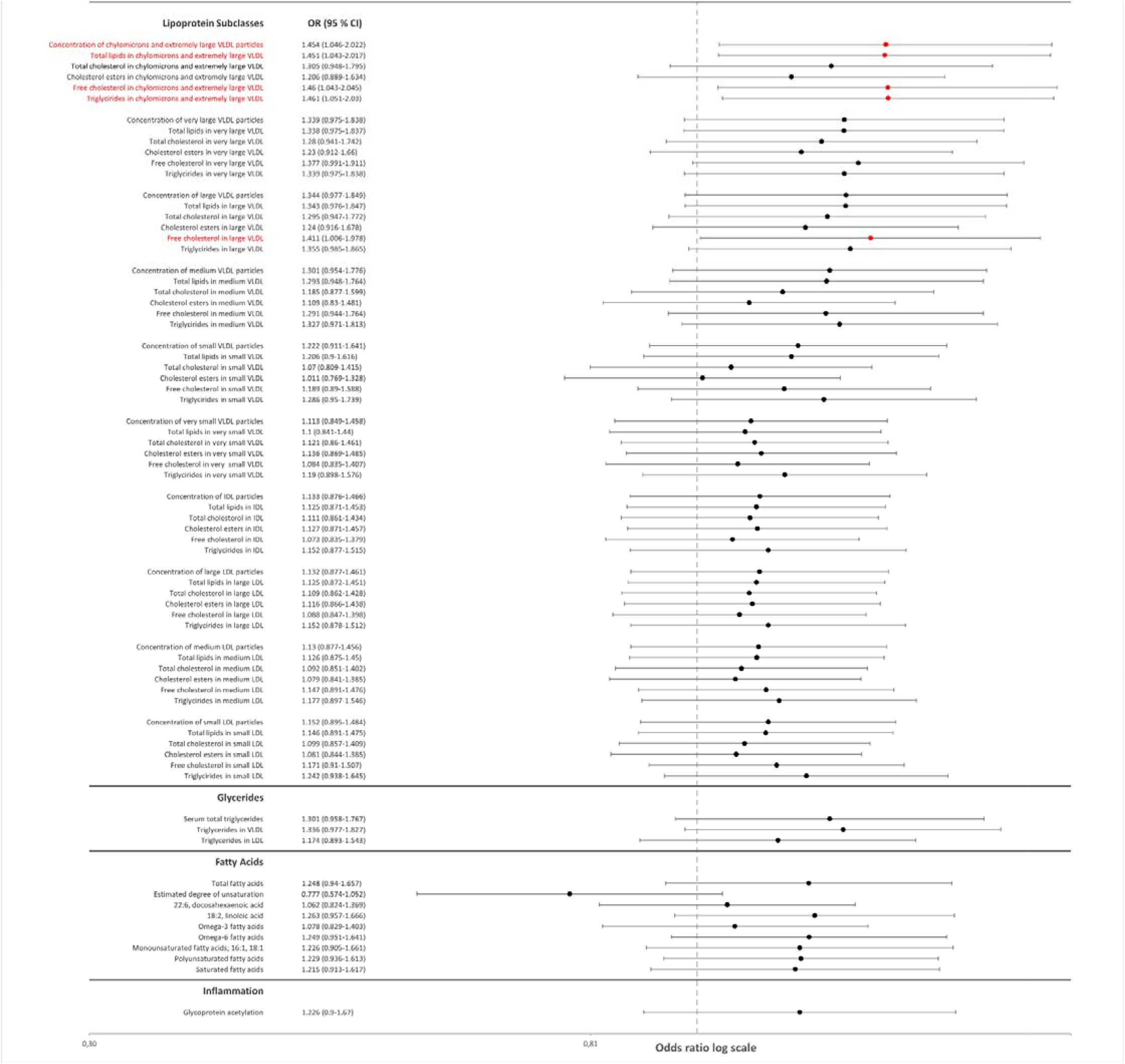
Results of the conditional logistic regression analysis of the 73 metabolic markers The measures highlighted in red were statistically significant in the univariate analysis but not after correction of multiple testing.

As a sensitivity analysis, we repeated the analysis in the subgroups of maternal BMI over 25 and 30 only. Although not statistically significant, the odds ratios (OR) of the metabolic variables were higher in these subgroups compared to the original population. The mean OR in the group with a BMI over 25 was 1.3 (N= 111, total) and in the group with a BMI over 30 was 2.1 (N= 42, total). BMI was included as a covariate in the analysis.

## Discussion

This study of maternal first-trimester metabolomic markers found that higher concentrations of four subtypes of extremely large VLDL particles and one subtype of large VLDL particles were observed in mothers with TGA in their offspring compared with mothers with a healthy child. Although it did not reach statistical significance after multiple testing correction, the trend in our BMI-matched cohort could indicate that the maternal early-pregnancy lipid profile, especially an extremely high VLDL composition, is associated with the likelihood of TGA development. The increasing OR in subgroups with higher BMI potentially signals a stronger association between TGA and metabolic markers in obese women.

Our aim was to identify early pregnancy biomarkers contributing to the association between maternal metabolic risk factors and CHD in the offspring. As the connection between CHD and higher maternal BMI has already been established in previous studies(11–13), the present analysis matched the groups with higher BMI. The proportion of overweight and obese women in this study was 38 %, which is similar to the general Finnish population where 35 % of women aged 18–29 years and 44.2 % aged 30–39 years have a BMI over 25 (20).

Moreover, the prevalence rates of diabetes and GDM were similar in both groups. Thus, our approach allowed us to identify potential pathophysiological mechanisms beyond obesity and diabetes.

Higher early-pregnancy metabolic markers, such as increased concentrations of VLDL particles in several sizes and of small and medium sized HDL particles, have been demonstrated in women who develop GDM in later pregnancy compared to non-diabetic women (21). In obese mothers with GDM, elevated lipids and lipoprotein constituents in VLDL subclasses, and greater triacylglycerol enrichment across lipoprotein particles, among other differences, were detected at least 10 weeks prior to the diagnosis of GDM (22).

Moreover, a meta-analysis concluded that triglycerides are remarkably elevated in women with GDM across the entire pregnancy compared with women without increased insulin resistance (23). These previous findings emphasise the connection between an abnormal early-pregnancy metabolomic profile and the later development of GDM. However, as an opposite finding, a recent study discovered that maternal total lipid intake was inversely related to the risk for double-inlet ventricle (DIV). In this study, maternal diabetes and insulin use, nevertheless, increased the risk for DIV. (24) Our finding that the subtypes of extremely large VLDL particles tended to be elevated in mothers of TGA infants points towards an abnormal lipid metabolism being a potential risk factor for the developing cardiac structures.

Few studies have addressed early pregnancy serum metabolomics and risk for CHD in the offspring. One recent study demonstrated that the maternal lipid profile of high TG, Apo-A1, and TC/HDL-C levels in early pregnancy were associated with an increased risk for CHD in the offspring, including both mild and severe cases where ventricular septal defects formed the largest CHD group (25). Another study of the maternal lipid profile 16 months after the pregnancy showed higher total cholesterol, LDL and apolipoprotein B among mothers with a child with CHD compared to controls. However, as the samples were collected after the pregnancy, definitive conclusions of the causal associations cannot be made. (26)

Since the diagnosis of CHD is made at the earliest during the late second trimester, third trimester, or postnatally, examining early pregnancy maternal biomarkers in CHD pregnancies poses a challenge. Although CHD is the most common congenital malformation, the incidence remains rather low, and tens or hundreds of thousands of samples are needed to achieve sufficient sample sizes for screening studies. Our approach, which used screening samples collected for other purposes, allowed us to retrospectively pick samples from the pregnancies of interest collected during several years. However, the collection method was suboptimal for the NMR analysis, and some markers, such as glucose metabolism markers, had to be excluded, as they showed instability when compared to the standards of the method provider.

The molecular mechanisms behind maternal metabolic status and the development of CHD in the offspring remain largely unknown. Excess abdominal adipose tissue both subcutaneously and viscerally is correlated with circulating inflammatory biomarkers and oxidative stress (27). Biomarkers of oxidative stress, such as concentrations of homocysteine and oxidized glutathione involved in the transsulfuration pathway, have been shown to be significantly higher in women with pregnancies affected by congenital heart defects than in women with normal pregnancies (28). Oxidative stress was not studied in our cohort but we investigated a circulatory inflammatory marker, GlycA. GlycA is a novel measure of low-grade inflammation consisting of a complex heterogeneous NMR signal. The main acute-phase protein contributors to the serum GlycA signal are α_1_-acid glycoprotein, haptoglobin, α1 - antitrypsin, α_1_-antichymotrypsin, and transferrin. (29) In a recent study, GlycA was found to predict GDM during early pregnancy in overweight and obese women (21). In another study with a population of 26,539 women followed for 16 years, GlycA levels at the baseline correlated with the risk of diabetes. This finding potentially suggests the role of protein glycans and subclinical inflammation in the development of diabetes. (30) We found no statistically significant difference between cases and controls in the GlycA levels.

TGA infants had significantly lower head circumference as compared to the controls even when adjusted for GA. In a systematic review of 28 studies, 26 studies found an association between congenital heart defects and measures of reduced prenatal brain growth. Hypoplastic left heart syndrome (HLHS), tetralogy of Fallot (TOF), and TGA, were reported to show the strongest evidence of association. (31) In a large nationwide cohort with 924,422 live-born Danish singletons, many subtypes of CHD were associated with smaller HC, but only neonates born with TGA had a disproportionately smaller HC also relative to their birth weight (32). Thus the finding that HC is smaller in TGA children is in line with previous studies.

The strengths of our study included the use of national registries that are mandatory and of high quality (33,34). The TGA diagnoses were verified from patient records. The samples were collected during the first trimester of pregnancy, representing the circumstances during cardiac morphogenesis (35), making our approach relevant from a developmental view. A major weakness of our study was that due to sample handling procedures, we were unable to analyse the exact blood glucose levels and metabolic markers associated with it. However, the prevalence of diabetes was similar in both groups, as was the proportion of OGTTs performed, so it is likely that these would not act as confounding factors in the comparisons. Another weakness is the study sample size, which, although it is a national cohort, is small. This diminishes the statistical power, especially in the subgroups of a BMI over 25 and 30. In addition, because of the sensitivity of the analysis process used on the metabolic samples, we were forced to exclude outliers potentially affected by the process. Despite these weaknesses, our results warrant further studies on this subject with larger patient cohorts.

### Conclusion and clinical relevance

Our study indicates that unfavourable lipid profiles of expectant mothers may pose a risk factor for CHD in their offspring. Although not statistically significant, the higher association trends in women with a BMI over 25 and 30 indicate a dose response effect for the risk.

Further studies with larger study samples are needed especially in women with a BMI over 25 and 30 to establish definite associations between metabolic markers and risk for congenital heart defects in their offspring.

## Data Availability

All data produced in the present study are available upon reasonable request to the authors.

## Acknowledgements

We thank Tero Vahlberg for his help with the statistics.

